# Short-term stability of diluted treprostinil sodium for subcutaneous administration

**DOI:** 10.1101/2024.04.19.24306042

**Authors:** Margaret R. Sketch, Daniel Sauerstrom, Allison Marie Lange, Jane Hall Diehl

**Author notes:** **Corresponding author:** Name: Jane Diehl, RPh, MS, MBA, Address: 55 TW Alexander Dr, Durham, NC 27709. **Funding:** United Therapeutics Corporation supported the research and manuscript development for this work.

## Abstract

**Background:** The Remunity Pump for Remodulin (treprostinil) Injection is an infusion system indicated for use with subcutaneous treprostinil.

**Objective:** The objective was to assess the stability of diluted treprostinil sodium stored in Remunity pump cassettes.

**Methods:** Treprostinil’s chemical and physical stability were assessed over a 76-hour period at 40 degrees Celsius and 75% relative humidity after dilution to 0.01 mg/mL in sterile water for injection and 0.9% sodium chloride. Analyses were conducted via visual inspection, pH measurement, and a stability-indicating high-performance liquid chromatographic assay.

**Results:** At all time points, the treprostinil samples were consistently clear, colorless, and free of visible particulate matter, with pH levels 6.0-7.0. All assays were within 90.0%-110.0% of the label claim, with minimal variation.

**Conclusions:** Treprostinil diluted to 0.01 mg/mL was stable in Remunity pump cassettes for up to 76 hours at elevated temperature and humidity. No notable changes from label claim concentration values were observed.

## Introduction

Treprostinil sodium is a chemically stable, pH-neutral analogue of prostacyclin, a potent vasodilator that inhibits platelet aggregation. Treprostinil injection (Remodulin)^a^ was approved in the United States in 2002 for the treatment of pulmonary arterial hypertension (PAH) Group 1 to diminish symptoms associated with exercise.^1^ Treprostinil injection is administered as a continuous infusion by either subcutaneous (SC) or intravenous (IV) routes, with SC being the preferred mode of administration.^1,2^ The calculated flow rate is based on patient’s dose (ng/kg/min), weight (kg), and treprostinil concentration (mg/mL).^1^

Subcutaneous administration of treprostinil injection requires continuous infusion using a catheter and infusion pump designed for SC drug delivery. The infusion pump should be adjustable to approximately 0.002 mL/hour with alarms to indicate occlusion/no delivery, low battery, programming errors, and motor malfunctions. Additionally, the device must have delivery accuracy of ±6% or better, be positive-pressure-driven, and have a reservoir made of polyvinyl chloride, polypropylene, or glass. Alternatively, patients may use an infusion pump cleared for use with treprostinil injection.^1^ The Remunity Pump for Remodulin (treprostinil) Injection^b^ is the first infusion system indicated for use with SC Remodulin. This pump was cleared in December 2020 by the US Food and Drug Administration (FDA) for continuous SC delivery of Remodulin in adult patients ages 22 years and older.^3^

Remunity pump cassettes are designed for single use and contain Remodulin that can be delivered SC over 3 days (72 hours) via the Remunity pump. These cassettes are either pre-filled by a specialty pharmacy or directly by the patient or caregiver.^4^ The minimum flow rate of 16 μL/hour has incurred some challenges for lower-weight patients utilizing the Remunity pump. One solution to address this is to evaluate using diluted treprostinil injection in the Remunity pump, allowing for delivery of a lower dose. This study sought to assess the stability of diluted treprostinil injection using two common diluents, sterile water for injection (SWFI) or 0.9% sodium chloride injection (normal saline, NS), and subsequent storage in Remunity pump cassettes. This study applied previously validated and published methods to determine the short-term physical and chemical stability of treprostinil injection for SC administration when combined with these diluents.^2^ Three evaluations were conducted to assess solution stability: physical liquid appearance; pH testing; and an assay using high-performance liquid chromatography (HPLC). HPLC is an analytical laboratory technique used to separate components of a mixture based on their chemical properties to provide information on the abundance of each component. In this study, HPLC was used to quantitate treprostinil label claim in treprostinil injection samples.

## Methods

### Solution preparation and storage

For this analysis, commercially available treprostinil sodium for injection (Remodulin)^c^ (1 mg/mL) was used to calculate and confirm the label claim. It was diluted in either United States Pharmacopeia (USP) SWFI^d^ or NS^e^ to 0.01 mg/mL (10,000 ng/mL), considered the label amount for the formulation, by adding treprostinil to a 100-mL volumetric flask and diluting to volume. Pending transfer to syringes, the solutions in flasks were stored at approximately 5°C. A total of 60 syringes^f^ (30 SWFI, 30 NS) were each filled with approximately 3 mL of the diluted treprostinil solution (0.01 mg/mL). Syringe contents were transferred to individual Remunity pump cassettes^g^ (3 mL capacity). As shown in the Supplemental Digital Content 1 (Table showing storage conditions and count of the stability units), 30 cassettes (15 SWFI and 15 NS) were intended for sample analysis. The remaining 30 cassettes (15 SWFI and 15 NS) were held in reserve.

To fill the Remunity pump cassettes (Figure 1), it was verified that the Luer Lock on the tubing was securely closed. Next, a 26-gauge x 5/8” needle was inserted in the cassette filling aid port, past both rubber membranes, and the full volume of the liquid was carefully discharged into the cassette. Subsequently, the cassette was examined for leaks. If a leak was observed, the cassette was discarded, and a new cassette was prepared. All cassette units were stored in a light-protected stability chamber at 40°C (±2°C) and 75% (±5%) relative humidity (RH) for a target of 76 hours (range 76-80 hours) with the black side of the cassette facing downwards.

**Figure 1.**
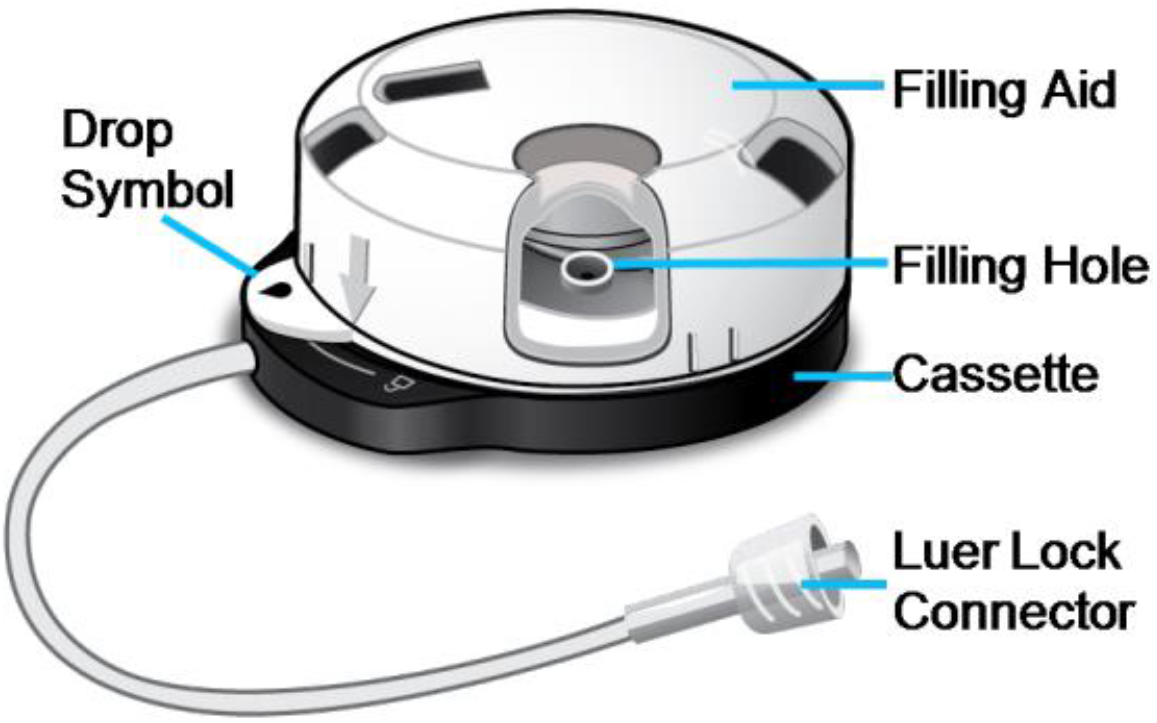
Remunity pump cassette.

### Testing protocols

Samples were evaluated for Remodulin assay content (% of label claim), physical appearance, and pH at time zero and the 28-, 52-, and 76-hour time points (t_0_, t_28_, t_52_, t_76_). All liquid obtained for analyses was evaluated as-is with no further dilutions, corresponding to a target treprostinil concentration of 0.01 mg/mL (10,000 ng/mL). For t_0_ evaluation, an aliquot for assay analysis and samples for visual analysis and pH determination were obtained from the 100 mL glass volumetric flasks in which the dilutions were performed.

At each time point subsequent to t_0_ (t_28_, t_52_ and t_76_), five Remunity pump cassettes for each diluent were removed from storage in the stability chamber and cassette contents were expelled into scintillation vials. To accomplish this, each cassette was activated by depressing the white casing, removing the Luer Lock, and rotating the unit to expel the liquid contents into an individual scintillation vial. Once emptied, the cassette was considered used and was discarded. Aliquots for assay determination were removed from three individual scintillation vials and transferred to an HPLC vial. The HPLC vials for assay determination were subsequently stored at approximately 5°C until final samples were collected at t_76_ and then analyzed in a single HPLC run.

A validated, stability-indicating HPLC method^h^ was used to confirm the label claim for each sample. A single label claim determination was made for each initial preparation of the sample solution (t_0_), while samples (n=3) were evaluated at each subsequent time point (t_28_, t_52,_ and t_76_).

HPLC^i^ instrumentation included a gradient pump,^j^ temperature-controlled autosampler^k^ at ambient temperature, an ultraviolet light detector^l^ set at a wavelength of 217 nm, and a temperature-controlled column heater^m^ set at ambient temperature. Sample analyses was conducted using a column^n^ measuring 4.6 mm x 250 mm, with a 5 μm particle size. The injection volume was 20 μL with a flow rate of 2.0 mL/min, a sampling rate of 5 Hz, and a run time of 42 minutes. For mobile phase A, a solution of 0.1% trifluoroacetic acid (TFA) in 60:40 water:acetonitrile was used. For mobile phase B, a solution of 0.1% TFA in 22:78 water:acetonitrile was used. Between samples, a 20:80 water:acetonitrile solution was used for needle wash. Pre-determined criteria to demonstrate instrument system suitability were met prior to sample evaluation.

At each time point following assay sample collection, the remaining solutions from the five scintillation vials were combined into a composite sample for each diluent for visual appearance evaluation and pH determination immediately following sample preparation. Product appearance was visually^o^ inspected for color, form, and the presence of visible particles; this was conducted under ambient laboratory lighting against a white background, while rotating the container. Visual inspection results were documented. The pH testing was conducted using USP <791> standardized methods, and results were documented.

All testing was conducted at Cambrex Corporation, Durham NC.

## Results

Table 1 summarizes HPLC results for the 0.01 mg/mL treprostinil samples diluted with SWFI- and NS, with results shown for both individual and averaged samples. At all time points, the individual and average samples values were within 90.0% to 110.0% of the label claim, meeting typical requirements for drug product stability,^5^ and showed minimal variation (~3%).

**Table 1.** PH Assessment and Label Claim Determination Results for Diluted Treprostinil (0.01 mg/mL) over 76 Hours.

| Formulation | pH | Preparation | Label Claim (%) | Average (%) |
| --- | --- | --- | --- | --- |
| SWFI, t<sub>0</sub> | 6.59 | NA | 102.8 | NA |
| NS, t<sub>0</sub> | 6.18 | NA | 101.8 | NA |
| SWFI, t<sub>28</sub> | 6.97 | 1 | 104.5 | 103.9 |
|  |  | 2 | 104.3 |  |
|  |  | 3 | 102.9 |  |
| NS, t<sub>28</sub> | 6.57 | 1 | 102.8 | 102.8 |
|  |  | 2 | 101.9 |  |
|  |  | 3 | 103.7 |  |
| SWFI, t<sub>52</sub> | 6.98 | 1 | 103.6 | 103.6 |
|  |  | 2 | 103.3 |  |
|  |  | 3 | 103.8 |  |
| NS, t<sub>52</sub> | 6.57 | 1 | 103.2 | 103.1 |
|  |  | 2 | 102.8 |  |
|  |  | 3 | 103.3 |  |
| SWFI, t<sub>76</sub> | 6.93 | 1 | 102.4 | 103.0 |
|  |  | 2 | 103.3 |  |
|  |  | 3 | 103.2 |  |
| NS, t<sub>76</sub> | 6.50 | 1 | 104.9 | 103.8 |
|  |  | 2 | 102.3 |  |
|  |  | 3 | 104.2 |  |

Abbreviations: NA, not applicable; NS, normal saline; SWFI, sterile water for injection; t, time (hours).

As shown in Supplemental Digital Content Table 2, (which summarizes appearance results for the SWFI and NS at t_0_, t_28_, t_52,_ and t_76_), the treprostinil samples were consistently clear and colorless on visual evaluation and free of visible particulate matter at all time points. All samples also had a measured pH between 6.0 and 7.0. At t_0_, the pH of dilute treprostinil in SWFI and NS were 6.59 and 6.18, respectively (Table 1). When tested at later time points (t_28_, t_52,_ and t_76_), the pH of all samples with both diluents showed a slight increase, ranging between 6.93 and 6.98 for the SWFI solution, and between 6.50 to 6.57 for the NS solution.

## Discussion

This study evaluated the chemical and physical stability of treprostinil sodium for injection diluted to 0.01 mg/mL in SWFI and in NS in Remunity pump cassettes for up to 76 hours at 40°C and 75% RH. Results showed that both diluted treprostinil solutions were stable at all time points up to 76 hours when stored at elevated temperature and humidity. No visible changes were apparent in the solutions on appearance testing, and the pH levels of all samples remained between 6 and 7. These findings are consistent with prior research showing that treprostinil injection maintained stability when combined with common diluents, then stored and subsequently delivered via an IV infusion pump.^1,2^ There are several clinical scenarios in which these data may be applicable. For example, treprostinil sodium for injection diluted to 0.01 mg/mL in Remunity pump cassettes may be necessary for lower-weight patients requiring a dose that would be below the minimum flow rate of the Remunity pump. As another example, institutions may be able to start Remunity in an inpatient setting, as the lower concentrations allow a lower initial dose and subsequent up-titration.

The stability analyses used visual inspections, pH measurements, and assays. Even though the method used for assaying the 0.01 mg/mL concentration is better used for formulations as low as 1.0 mg/ml, the results for the diluted treprostinil solutions were above the limit of quantitation, with the focused intent of demonstrating stability over time when stored in the Remunity cartridge. The detection of impurities or degradation products was not in the scope of this study, and without further evaluation it is unclear if the method used would be able to detect impurities at the Remodulin dilution level evaluated.

## Conclusions

Treprostinil diluted with either SWFI or NS to as low as 0.01 mg/mL was stable in Remunity pump cassettes for up to 76 hours at elevated temperature and humidity. No significant changes from label claim values were observed for either formulation at any time point. Likewise, the solution’s appearance and pH were largely unaffected by dilution and subsequent stability evaluation.

## Data Availability

All data produced in the present study are available upon reasonable request to the authors

## Acknowledgements

The authors would like to thank Stephanie Hwang, PharmD for her support during development of this manuscript. Caitlin Rothermel, MPH provided editorial assistance.

## What’s New

○ Treprostinil diluted with sterile water for injection or normal saline to 0.01 mg/mL was stable in Remunity pump cassettes for up to 76 hours at elevated temperature and humidity with no significant differences from label claim at any time point in appearance, pH, or stability.
○ Diluted treprostinil may be useful for lower-weight patients utilizing the Remunity pump because the pump flow rate does not allow for low-dose delivery.
○ Diluted treprostinil for Remunity Pump administration may enable therapy initiation in lower-weight patients, including hospitalized patients, as the lower concentrations allow for a lower initial dose and subsequent up-titration.

## Figure captions

**Supplemental Digital Content Table 1.** Stability Unit (Remunity Pump Cassette) Requirements: Storage Conditions and Count.

| Formulation | Time Point (hours) | Stability Storage Conditions | Units Required for Testing (cassettes) | Units Required for Reserve Storage (cassettes) | Total Units (cassettes) |
| --- | --- | --- | --- | --- | --- |
| Diluted in SWFI | t<sub>28</sub> | 40°C/75% RH | 5 | 5 | 10 |
| Diluted in NS | t<sub>28</sub> | 40°C/75% RH | 5 | 5 | 10 |
| Diluted in SWFI | t<sub>52</sub> | 40°C/75% RH | 5 | 5 | 10 |
| Diluted in NS | t<sub>52</sub> | 40°C/75% RH | 5 | 5 | 10 |
| Diluted in SWFI | t<sub>76</sub> | 40°C/75% RH | 5 | 5 | 10 |
| Diluted in NS | t<sub>76</sub> | 40°C/75% RH | 5 | 5 | 10 |

Abbreviations: NS, normal saline; RH, relative humidity; SWFI, sterile water for injection; t, time (hours).

**Supplemental Digital Content Table 2.** Appearance Testing Results for Diluted Treprostinil (0.01 mg/mL) over 76 Hours.

| Formulation | Time Point | Appearance |
| --- | --- | --- |
| SWFI | t<sub>0</sub> | Clear, colorless liquid, free of visible particulates |
| NS | t<sub>0</sub> |  |
| SWFI | t<sub>28</sub> |  |
| NS | t<sub>28</sub> |  |
| SWFI | t<sub>52</sub> |  |
| NS | t<sub>52</sub> |  |
| SWFI | t<sub>76</sub> |  |
| NS | t<sub>76</sub> |  |

Abbreviations: NS, normal saline; SWFI, sterile water for injection; t, time (hours).

## Footnotes

a United Therapeutics, Silver Spring, MD

b United Therapeutics, Silver Spring, MD

c United Therapeutics, UT-15 Reference Standard, Lot D-1013-147

d Corning, Lot 31222012

e KD Medical, lot 010623-01

f BD 3 mL Sub-Q Syringe with Luer-Lok, 26 G x 5/8 (0.45 mm x 16 mm), Part number 309587

g UT, DKPI-11022-001

h Cambrex Corporation, DUR-QCT-1897-TM

i Agilent 1290 Infinity II UHPLC

j Agilent 1290 Infinity II Flexible Pump, model G7104A

k Agilent 1290 Infinity II Vial sampler, model G7129B

l Agilent 1260 Diode Array Detector (DAD), model G7117C

m Agilent 1260 Infinity Multicolumn Thermostat (MCT), model G7116A

n YMC America, YMC Pack ODS-AQ C18

° Cambrex Corporation, DUR-QCT-0106-TM

